# The effect of access to safe Water, Sanitation, and Hygiene (WASH) facilities on Child Growth Failure among children 6/59 months in Ecuador applying a random intercept multilevel model using cross-sectional ENSANUT 2018 data

**DOI:** 10.1101/2022.03.23.22272829

**Authors:** Buizza Cristiano

## Abstract

1.

**Background:** Child Growth Failure - measured as stunting, wasting and underweight - is still an important public health issue affecting 23.1% of children. Typically, literature focused on unproper dietary habits, but living in an unhealthy environment unable to prevent pathogens is another fundamental cause.

**Objective:** To estimate the association between access to safe water, sanitation, and hygiene (WASH) facilities and risk for Child Growth Failure in under-5 children in Ecuador **Design:** Cross-sectional multilevel study using the Ecuadorian National Survey on Health and Nutrition (ENSANUT) 2018.

**Participants:** 17,688 children 6/59 months residing in 15,382 households.

**Main outcome measures:** Association between the access to safely managed WASH factors and the probability of suffering from: a) stunting, b) underweight and c) wasting; controlling for child’s sex, child’s age, antenatal care visits, pre-term delivery, assumption of micronutrient during the pregnancy, mother’s age, mother’s education, mother’s height, mother’s ethnic group, area and region of residence, the number of people living at home and the family per capita income. The final model is a two-levels random intercept logistic regression focused on the risk of suffering from stunting and underweight. Multilevel logistic models were applied for both the unadjusted and the adjusted estimates. The average marginal effects with the 95% confidence interval and p-value are estimated.

**Results:** A safely managed sanitation system showed the strongest protective effect on underweight (−38.1%, 95% CI -16.9% and -59.4%) and stunting (−14.9%, 95% CI -4.7% and -25.1%). Important protective effects against stunting are observed also thanks to safely managed drinking water (−10.9%, 95% CI -0.1% and -21.6%) and applying proper hygiene habits (−9.5%, 95% CI -0.1% and -19.0%). A potential protective effect on underweight was observed also for proper hygiene habits although a wide confidence interval. No effect was observed between safely managed drinking water and the risk of being underweight. Women are characterized by an important lower risk for both stunting and underweight. Stunting is lower after the 24^th^ month of life while no change is observed for underweight. Pre-term deliveries showed a strong growth of the risk for both stunting and underweight, while antenatal care visits significantly reduced the risk of both stunting and underweight as a high mother’s education. Genetical (mother’s height) and cultural (mother’s ethnic group) aspects play an important role with the highest risk for stunting among Indigenous (+32.5%, 95% CI 19.6%-45.4%) and the lowest among Afro-Ecuadorian children (−20.7%, 95% CI 2.0% and -43.4%). Each additional centimetre of the mother’s height from the average value (154.2 cm) reduces the risk for the child of suffering from both stunting (−6.1%, 95% CI -5.3% and -6.7%) and underweight (−5.1%, 95% CI -3.7% and -6.5%).

**Conclusions:** WASH factors play a fundamental role to prevent undernutrition, especially chronic undernutrition (stunting). The study found potential biases due to the use of self-reported cross-sectional data. No data on dietary habits were available for this study which is a potential lack to be considered for the future.

## 2. INTRODUCTION

Child Growth Failure (CGF) defines a condition in which children are characterized by slower growth rates compared the appropriate growth velocity for the age and sex due to a muscle and fat mass lack associated with undernutrition. Anthropometric measures such as weight, height, mid-upper-arm circumference or skinfold thickness can be considered as proxies (more or less precise) of the muscle and fat mass lack. Due to logistical simplicity, weight and height (and derived measure such as body mass index and wasting) are the most common anthropometric values used especially to compare among countries [Black et al. 2013, Osgood-Zimmerman et al. 2018]. Stunting (low height for age), wasting (low height for weight) and underweight (low weight for age) are the three indicators stemming from height and weight. Despite a steadily decline during the last two decades, the prevalence of under-5 stunting, wasting and underweight still remains high especially in Southern and South-eastern Asia and in West, East and Middle Africa [UNICEF/WHO/World Bank 2021].

Stunting, otherwise defined as chronic undernutrition, is the product of poor nutrition and health conditions in-utero and during the early childhood. According to the most updated estimates [UNICEF/ WHO/WB 2021], in 2020 globally around 22% of children (approximately 149.2 million) suffered from chronic undernutrition. Although, Latin America and the Caribbean (LAC) shows a lower prevalence (11.3% compared to 30.7% of Africa and 21.8 of Asia), important differences can be observed at national and subnational level. In Ecuador, under-5 stunting is still a persistent public health concern with a national prevalence of 23.1%, reaching a peak of 28.7% in the rural areas of the country [INEC 2019].

A different situation is observed focusing on wasting (acute undernutrition) which is an extreme thinness for height. Ecuador, as in most of LAC countries, shows a low prevalence of wasting equal to 3.7%. Wasting (as well as stunting) is the result of both a poor nutrient intake or absorption, due to environmental enteric dysfunctions (EED) which can deteriorate the immunity system through systemic inflammations, and due to nutrient losses for infections such as diarrhoea [Keusch et al. 2013, Bourke et al. 2016, Owino et al. 2016, Komarulzaman et al. 2017, Luby et al. 2018, He et al. 2018, Yaya et al. 2018, Dagnew et al. 2019, Pickering et al. 2019, Solomon et al. 2020].

Stunting and wasting are two notions generated to differentiate among underweight due to a low weight in relation to the height and underweight due to a small height in relation to the age [Waterlow 1977]. So, although they are normally presented as a measure of alternative forms of undernutrition, these indicators are strictly related and caused (at least partially) by common causes [Briend et al. 2015]. Consequently, the purpose of this study is to find common factors able to explain the risk for suffering from stunting, wasting or underweight.

Although some children can suffer mutually of stunting and wasting, increasing even more the risk of death, the focus of this study will be the independent risk for each condition.

Additional to poor nutritional intake (in terms of quality, quantity and diversity), another important cause for CGF is living in an unhealthy environment unable to prevent diseases. Lack in the access to basic health, sanitation, water and hygiene services raises the risk of diarrhoea, environmental enteropathy and nematode infections which are direct causes of an augmented risk of morbidity in children, increasing the risk for suffering from CGF [Checkley et al. 2008, Dangour et al. 2013, Richard et al. 2013, Keusch et al. 2013, Owino et al. 2016]. Ending open defecation, a safe management of water and sanitation, and proper hygienic habits are important factors able to reduce the exposure of population to pathogens causing diarrhoea and environmental enteric disorders common in young children [Komarulzaman et al. 2017, Luby et al. 2018, He et al. 2018, Yaya et al. 2018, Dagnew et al. 2019, Pickering et al. 2019, Solomon et al. 2020].

Due to The Millennium Development Goals signed in 2000 and especially the Sustainable Development Goals (SDG) set up in 2015, the last two decades experimented a rise in interest in the association between improved water, sanitation and hygiene habits (WASH factors) and CGF. Extending the access to drinking water and improving sanitation and hygiene is a priority explicitly expressed in SDGs 6.1 and 6.2. Nevertheless, although a strong impact observed on the risk of suffering from enteric diseases or child mortality, inconsistent or no empirical evidence are encountered in most of the studies focused on WASH factors and CGF [Arnold et al. 2009, Langford et al. 2011, Bowen et al. 2012, Cameron et al. 2013, Dangour et al. 2013, Clasen et al. 2014, Luby et al. 2018, Headey and Palloni 2019, Pickering et al. 2019, Bekele et al. 2020].

Potential benefits are observed in Luby et al. (2018), where a cluster randomized controlled trial developed in Bangladesh found significant improvements in height-for-age, weight-for-age and weight-for-height z-scores thanks to WASH factors but just when combined with nutrition treatment. Similar outcomes are observed in Null et al. (2018) with a cluster randomized controlled trial developed in rural Kenya. The non-random trial developed by Fenn (2012) found a decrease of 12.1% in the prevalence of stunting between the treatment and the control group. Bekele et al. (2020), using a cross-sectional analysis in Ethiopia, found a significant reduction of 29% in stunting among children with access to improved sanitation with handwashing facilities and a significant reduction of 17% of underweight for children with access to improved handwashing. Creating a panel data of 442 regions derived from the Demographic and Health Surveys, Heady and Palloni (2019) found a slight reduction in child stunting associated with access to piped water into house.

An important reason for the inconsistency observed in the impact of WASH on stunting, wasting and underweight is due to the different designs applied [Headey and Palloni 2019]. While randomized control trials normally are not able to find significant association, especially due to the short duration of exposure to WASH factors, observational studies tend to observe a stronger and significant one. Important methodological limitations, which can generate bias in the estimations, are observed also for observational data. Normally, WASH exposures are not linked to any specific intervention and are potentially associated with contextual confounders that cannot be considered in the analysis such as local hygiene habits, economic development, historical infrastructural presence, population density, and so forth [Headey and Palloni 2019].

The significance or at least the magnitude of the association between WASH and CGF can change also depending on the definition of exposures [Gunther and Fink 2010, Headey and Palloni 2019]. In general, there is no aprioristic strategy to classify all the existing WASH services as safe or improved or basic according to the effect on child health, although it seems that moving from open defecation to any form of toilet is the primordial cut-off able to improve health [Gunter and Fink 2010]. But varying the cut-off point, it changes the observed impact. Headey and Palloni (2019) found that the effect of improved water on child stunting rose if instead of the traditional classification formulated by Joint Monitoring Program (JMP) developed by World Health Organization and UNICEF [UNICEF/WHO 2019], a classification that disaggregates water system based on technological aspects is used^2^ [Gunther and Fink 2010].

Most of the studies focused on the association between WASH factors and child growth performance have been conducted in rural settings of Africa and Asia, while a gap on this topic can be observed in South America and especially the Andean region – Colombia, Ecuador, Peru and Bolivia – although an important prevalence of both WASH deficiencies and physical growth lack. Therefore, an analysis using the Ecuadorian National Survey on Health and Nutrition (ENSANUT) 2018 is applied to answer the following research questions:

1. Do the children living in households with a safely managed water system have a lower risk (or odds) for a) stunting, b) wasting and c) underweight compared with children living in household with no safe water system?
2. Do the children living in households with safely managed sanitation system have a lower risk (or odds) for a) stunting, b) wasting and c) underweight compared with children living in household with no adequate sanitation system?
3. Do the children living in households where people adopt appropriate hygiene habits have a lower risk (or odds) for a) stunting, b) wasting and c) underweight compared with children living in household in which people does not adopt appropriate hygiene habits?

## 3. METHODS

### 3.1 Data and sample power

This study used the Ecuadorian National Survey on Health and Nutrition (ENSANUT) 2018, a two-stage cross-sectional survey specifically developed to study under-5 stunting phenomena in Ecuador among others. The first stage was based on the selection of geographical units (primary sampling units - PSU) and within each PSU households have been selected. Data have been designed to be representative at national, urban, rural, as well as provincial (24) level and to collect information on demographic, socioeconomic, nutritional, health-related, physical, mental and emotional development variables [INEC 2019]. Database was freely available from Ecuadorian National Institute of Statistics webpage and data have been anonymized using identifiers.

The number of households (clusters) available – both with and without children - was equal to 43,311 and the children under-5 were 21,588. All the children under-5 living within the selected households have been interviewed and measured. To minimize the risk for recall bias a subsample of the last three children under-5 born in the five-years period before the interview was used as a specific section of the survey has been developed for this group.

Due to the different habits of feeding, just children of six months or older were considered. Out of the potential number of observations (18,535), in 679 cases no information was observed for both height and weight (17,856). Additionally, 168 missing values were observed for the confounders applied in the study. So, the final sample was equal to 17,688 children with an age 6/59 months at the moment of the interview and data on the response variable residing in 15,382 households.

As described in the introductory paragraph, the last updated estimates, previously to ENSANUT 2018, denoted a national prevalence of under-5 stunting equal to 23.1% (0.231), a prevalence of wasting equal to 3.7% (0.037) and a prevalence of underweight equal to 5.2% (0.052). Consequently, the stunting - as requiring the biggest sample size - was used to define the minimum sample size required. As data were correlated and deriving from a complex survey, a design effect (DEFF) equal to 2 was assumed. A sample able to produce highly precise estimates, with a confidence interval no wider than +/- 0.01 (considering the central parameter, a confidence interval between 22.1% and 24.1%), a *z* equal to 1.96 and therefore a standard error “*e*” equal to 0.005 was assumed. Individual and Households response rates (*rri, rrh*) were expected to be 95% (0.95).

As suggested in Kirkwood and Sterne (2003), formula adopted to estimate the sample size required for that level of precision has been: 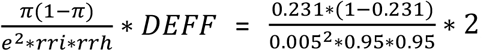, which was equal to 15,747 observations, corroborating that the available sample of 17,688 children 6/59 months was big enough to guarantee the desired precision.

### 3.2 Outcomes

The condition of stunting, wasting and underweight was based on the position of each child along the standard normal distribution of the reference group. The z-score is the dimensionless quantity which defines, for a measure of interest (weight, height, BMI), the individual distance of a specific person from the population median of the healthy group of reference, normally of the same sex and age [Wang and Chen 2012]. The distance is expressed in terms of standard deviations (SD). The reference population derives from the WHO 2006 Child Growth Standards [WHO 2006]

According to Wang and Chen (2012), a child is stunted when it is too short for his or her age which happens if the height-for-age z-score is less than -2SD from the mean value observed in the reference population. Additionally, a child is wasted and/or underweight if the weight-for-height and the weight-for-age z-scores are lower than - 2SD compared to the WHO healthy reference population.

For each observation, the weight and height were measured three times. After having calculated the distance between each measure, the most distant value was eliminated and the average between the other two measures was used to define the weight and the height of each person. Implausible values according to WHO standards have been eliminated^3^.

### 3.3 Exposures

As previously observed, different classifications of the exposures can produce different outcomes. The Joint Monitoring Program (JMP) [UNICEF/WHO 2019] classifies water services as “safely managed” if: a) accessible to dwelling, yard or plot, b) available at least 12 hours per day and c) free from E.coli, arsenic and fluoride. Piped water on the premises tends to be less contaminated especially if connected to public system due to quality controls that periodically has to be conducted. However, the quality of the water can be affected by transportation or by the storage strategy adopted at the household level which can generate a recontamination at the point of use despite a clean water at the source [UNICEF/WHO 2019].

Similar concerns arise for the sanitation system. According to the JMP, sanitary system can be classified as “safely managed” if facilities are not shared with other households and the excreta are: a) eliminated through the sewer lines, or b) emptied from septic tanks and latrine pits, or c) treated in situ in septic tanks or in latrines with appropriate strategy [UNICEF/WHO 2019]. However, several issues may appear due to poor-design, damage or flooding which can compromise a safe management of excreta. Additionally, a safe management of excreta requires a constant cleaning and treatment of septic tanks and latrines which may not be the case. A survey conducted in Ecuador in 2017 on septic tank and pit latrines found that in 87% of cases they do not effectively contain or treat excreta and that in 89% of cases they had never been emptied^4^, exposing children to pathogens [UNICEF /WHO 2019].

Considering all the issues observed, several studies [Gunter and Fink 2010, Headey and Palloni 2019] suggested the application of different strategies to classify water and sanitation systems. In the case of Headey and Palloni (2019), authors disaggregated JMP improved water system classification distinguishing among: a) piped to home, b) piped to other (public tap, standpipe and neighbour) and c) other improvement such as tube wells, boreholes and protected wells/springs. Gunter and Fink (2010) proposed an alternative classification based on technological aspects in which “water services” are categorized in the groups: a) “piped” which includes piped water into dwelling, yard or plot and public tap or standpipes, b) “well” which contains the cases when water is provided through all the possible types of wells, boreholes and springs, and c) “surface” when water is obtained directly to surface water. Also “sanitation services” are labelled in three categories: a) “flush” which includes shared and unshared flush toilets with sewer systems or septic tanks, b) “latrine” with all the types of latrines, and c) “open” for open defecation. Additionally, the authors suggest the possibility to differentiate water and sanitation systems based on private or public access [Gunter and Fink 2010].

Combining the strategies observed in literature, for the current study “water systems” have been classified as safely managed if provided by a public or a private entity authorised to control and treat water against pathogens. So, this category included: a) drinking water piped through a public or private system both to the dwelling, yard, plot or public standposts and b) packaged water, including bottled water and sachet water. Delivered water, including tanker trucks and small carts, was not included in the category as normally provided by microenterprise, often operating with no specific license and lack in water controls and water treatment. Boreholes, tube wells, wells and spring, rainwater, surface water^5^ and not drinking water in general were classified as “unimproved” as at higher risk of no proper treatment in case of pathogens contamination.

Focusing on the sanitation system, due all the issues previously stressed, just unshared flush toilets with sewer system has been considered as safely managed. All the other cases (shared toilets, septic tanks and latrines) were considered as “not safely managed”. Open defecation is a habit practically not existing in Ecuador, so the extremely small number of cases declaring this practice were included in the “not safely managed” category.

Access to appropriate hygiene was classified adopting the original JMP strategy which requires the joint presence at home of handwashing facilities, soap and water [UNICEF/WHO 2019]. All three exposures were at household level.

### 3.4 Control variables

Alternative explanations of child growth failure are: a) malnutrition (mainly associated with poverty) and failure to meet micronutrient requirements, b) diseases affecting bone and cartilage, nervous, circulatory, or gastrointestinal systems, c) genetics (familial short stature), d) hormonal deficiencies and e) other factors like intrauterine growth retardation or pre-term delivery [Martorell 1999, Branca and Ferrari 2002, Rivera 2003, Rogol et al. 2014, Sania et al. 2014].

Due to the strict association between health status and the quality of contextual and personal hygienic condition, all these items are potentially associated also with the access to WASH factors. The only exception is with genetics aspects which are considered in any case in this analysis due to the extreme importance for the response variable.

Malnutrition is probably the most common cause of growth failure and is usually associated with poverty. Unfortunately, the survey collects data on nutritional behaviors just for people up to 3 years which would generate a substantial amount of missing information if the variables would be included in the model. A question related with micronutrient intake during each pregnancy was used to measure the effect of potential micronutrient lack on child growth failure. Deficiencies of micronutrients, such as iron, magnesium and zinc may contribute to growth retardation reducing the intake of energy and protein. So, it was used a categorical variable distinguishing among pregnancy without receiving any type of micronutrient with pregnancies during which mothers were provided with iron or folic acid or both. A potential issue associated with this variable is that the intake of micronutrient could be the consequence of the detection of intrauterine growth restriction.

The level of wellbeing was measured collecting information on labor and/or other sources of income. Income data were summed and divided by the number of the component in each household to calculate the per capita family income. The variable related with poverty for unsatisfied basic needs was not used as some dimensions are related with the exposures and other confounders already included into the model.

For every person in the household, information is collected if they were sick during the last 30 days and the type of disease. As the data is derived from mothers’ opinions, the classification of diseases can be highly inaccurate. An alternative is the use of the question asking if the child suffered from diarrhea during the last 15 days before the interview. In this case concerns raise as it is related just with gastrointestinal disorders and it is a self-reported information with different mothers that could classify the same type of feces in a different way. An additional issue in this case is the presence of missing data^6^ (n=246, 1.4%, Table3). So, in this analysis none of the variables has been used.

Due to the large amount of missing data, both the variable associated with the weight at birth and the subjective question in which people were asked if the child at birth had a smaller, equal or bigger size than the other children have not been used (n=933, 4.5%, Table3). A self-reported variable in which mothers declare if delivery was pre-term or on-time was used.

Mother’s height is used as a proxy for genetics factors. The data used is the average of two remaining measurements after having eliminated the most different value for each mother. Height is an objective measure calculated by interviewer during the survey. Due to the importance of this factor, and to the fact that this variable is measured in an objective way, it was applied to analysis although the small amount of missing data observed (n=168, 0.9%, Table3). An analysis of missingness pattern was developed for this variable. Data on mother’s weight and the associated BMI were not used as values could be a consequence of the birth of a child. Hormonal deficiencies were not considered in the analysis due to a lack of information.

Other “a-priori” control variables – both at child and household level – were considered: child age and sex, mother’s age at delivery, and mother’s education as a better formal education is also associated with the knowledge for a better care. Mother’s ethnic group is used as a fixed effect for cultural and genetic aspects. Ethnic group is a self-declared condition which can be: a) Indigenous, b) Afro-Ecuadorian, c) Mestizos, d) Caucasian or e) Montubios. Sub-region, a combination of area of residence (urban or rural) and region (Coast, Sierra and Amazonia), is used as a fixed effect for contextual effects associated with the quality of the health services, which is different depending on the geographical area in which people reside, and the potential effect of the altitude. Additionally, to consider the effect of the access to health services the number of antenatal care visits is used. As the spread of pathogens is associated with the behaviour of all the family members, the number of people living in each household was used as a proxy for the number of people sharing the WASH facilities.

To facilitate the interpretation and to lessen potential collinearity due to the high number of variables used, all the continuous variables were centred. Centering is a simple statistical technique which does not change the slope between predictor and response variable but changes the interpretation of the intercept as after the transformation it represents the value assumed by dependent variable when the independent is equal to the reference value. In the case of per capita family income, values were centred around the median, so people earning below the central value can be considered as affected by relative poverty. In the case of antenatal care visits data were centred around four which is the minimum number of visits suggested by WHO [WHO 2016].

No post-delivery data were used as mothers with children suffering from detected CGF could go to the doctor at a higher rate generating a potential positive effect with the risk for CGF.

### 3.5 Statistical methods

As in each household all the under-5 children were measured, all the methods applied took into account the correlation among observations. Bivariate and a final multivariate model were used to examine the association between WASH factors and child growth failure (CGF) adjusting for potential confounders. For response variables and for confounders showing missing values it was investigated the missingness mechanism applying a multivariable logistic model. As children 0/5 months show different feeding habits, they were excluded from the analysis. As CGF is measured using three different indicators – stunting, wasting and underweight – three different blocks of models were developed. Both the initial unadjusted and the final adjusted multilevel models were estimated using software STATA15.

As the relationship between factors and the response variable is non-linear, the average marginal effects (AME), which denote the average effect of each parameter on the response variable after controlling for other variables, were estimated, joint with the 95% confidence intervals and p-values associated. As the AME is a different strategy from marginal effect at mean, it has been calculated also for continuous variables although already centred around the grand mean or around representative values. As stressed by Norton and Dowd (2018), to evaluate the effects of explanatory variables on binary response variable it is better using average marginal effects as odds ratio suffer from data dependency and cannot be used to compare different samples or different models with different sets of explanatory variables.

## 4 RESULTS

### 4.1 Descriptive analysis

As observed in figure1^7^, using the whole Ecuadorian under-5 population, the prevalence of each indicator (stunting, underweight and wasting) changes with the age. Stunting is the most important public health issue in Ecuador with an average prevalence equal to 25.7% among children recently born, which increases up to 29.2% among children between 6 and 23 months and a subsequent drop to 20.8% among the oldest children in the 24-59 months group^8^. Similarly downward trends with age are observed for both underweight and wasting. In the underweight category, the prevalence ranges from 10.2% among infants 0-5 months to 4.3% among children 24-59 months^9^; in the latter, the average prevalence of wasting ranges from 7.4% among infants 0-5 months to 2.8% among the last group (24-59 months)^10^. It is important to stress that confidence intervals of each age group are overlapped which means that potentially no significant changes are really observed while children get older.

**Figure 1.**
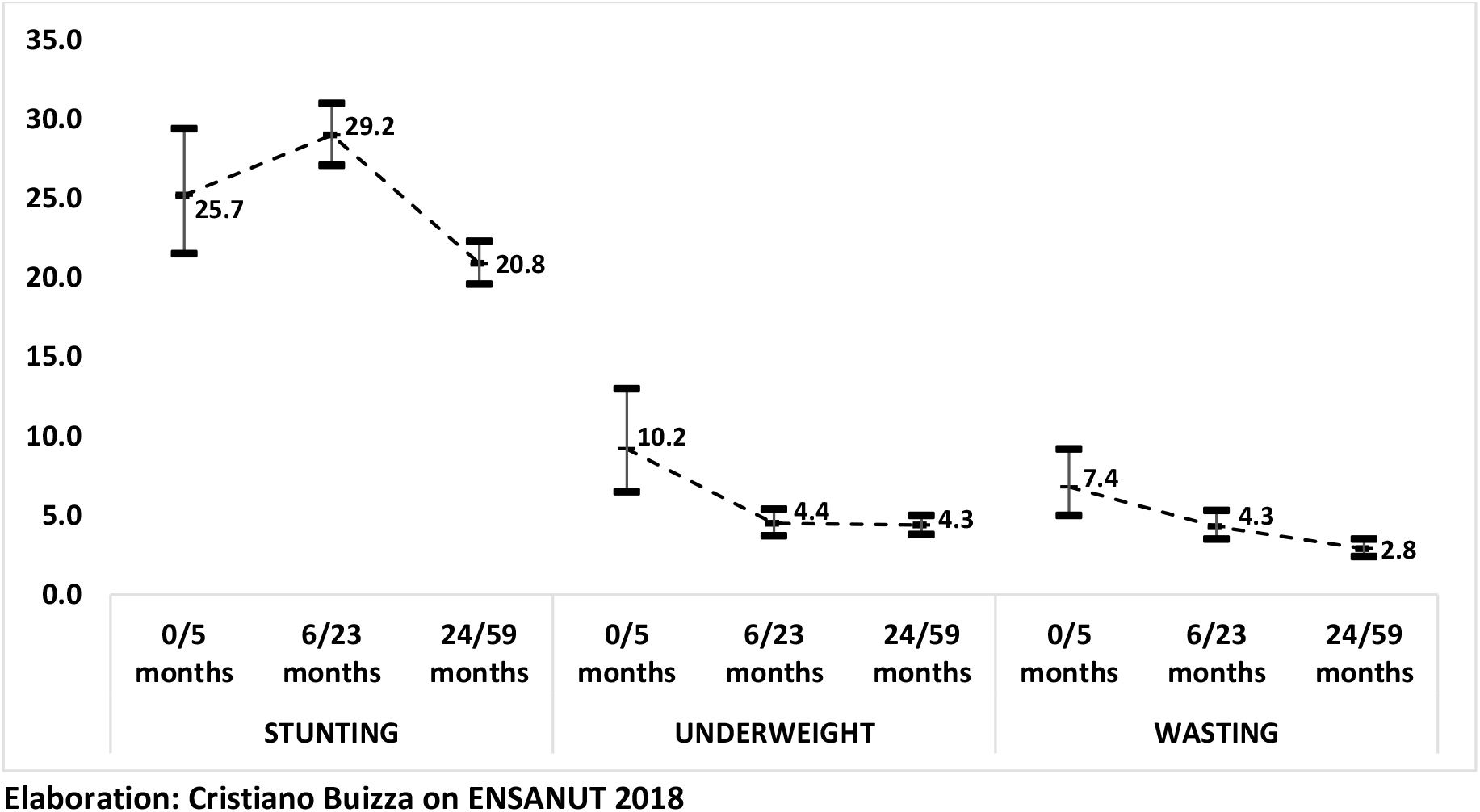
Prevalence of stunting, underweight and wasting among children by age. Ecuador, ENSANUT 2018.

As previously stressed, to increase the available information, the analysis is focused on the last three children of each mother. To correct for the effect of the different feeding habits, just children 6/59 months are used^11^. Table1 shows that 679 cases were missing on height and weight, so data are available for 17,856 children. On average, 23.6% [95% CI 22.5%-24.8%] of the children used for this analysis suffers from chronic undernutrition (stunting), 4.4% [95% CI 3.9%-4.9%] are affected by underweight and 3.3% [95% CI 2.9%-3.8%] shows a too low weight for the height (wasting or acute undernutrition).

**Table 1.**
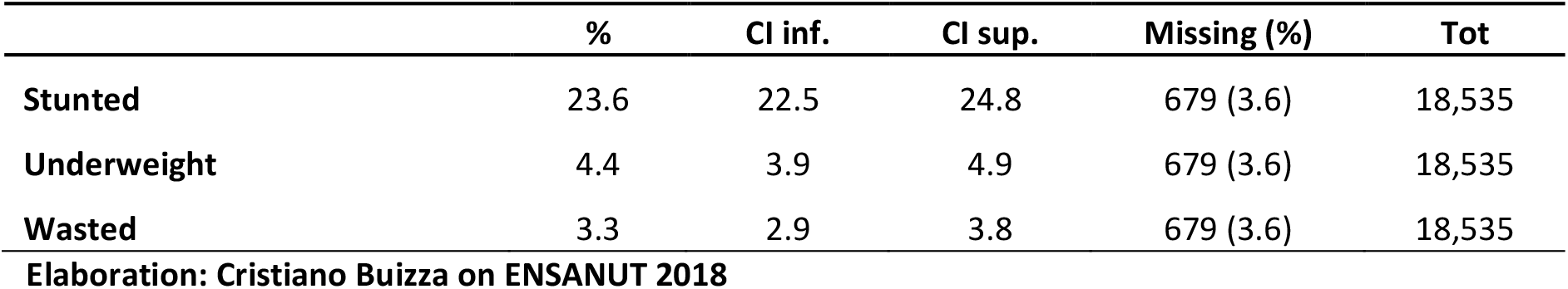
Percentage of children 6/59 months stunted, underweight and wasted. Ecuador, ENSANUT 2018.

Table2 shows the prevalence of children living in households equipped with safely managed water and sanitation services and with an adequate equipment for proper hygiene habits. As explained in the previous chapter, different strategies compared to Joint Monitoring Program (JMP) [UNICEF/WHO 2019] were applied, especially to classify sanitation system. In this case, more importance was applied to pathogens prevention. As almost 100% of people with septic tanks and pit latrines declares not cleaning properly the sanitation systems, all these cases are classified as “unsafe”. Consequently, just 53.9% of children [95% CI 52.1%-55.7%] live in households with safely managed sanitation system. For water systems and hygiene habits the percentage of children residing in households classified as safe is 75.9% and 74.5% respectively. No additional missing data are observed for exposures.

**Table 2.**
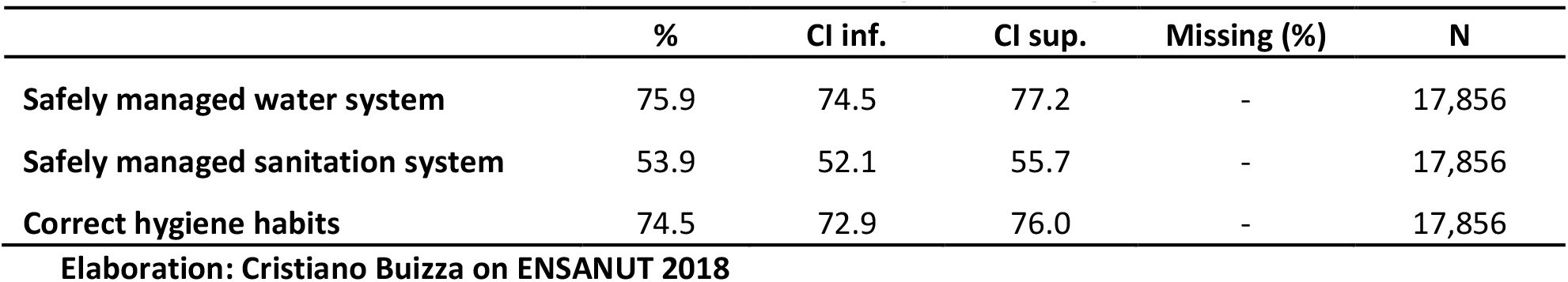
Percentage of children 6/59 months living in a household with safe WASH factors. Ecuador, ENSANUT 2018. Children with data on weight and height.

Table3 shows distribution of confounders among children (level 1) and household (level 2). It is observed a similar distribution of gender (51.4% male and 48.6% female), an average age of 32.4 months and an average number of antenatal care controls equal to 6.9. In 95.0% of pregnancies mothers received micronutrients during the pregnancy and for 11.9% of children the delivery was pre-term. Access to higher education is substantial as almost 65.5% of women had a secondary or tertiary level. Mothers who belong to the ethnic group of Mestizos are most of the sample with a percentage equal to 79.8%. The average height of the mothers who were in the sample was 154.2 cm which is below the expected value also for South America (160 cm) [WorldData]. The 67.9% of households are in the urban area of the country mostly situated in the Andean region (39.1%) and in the Coast region (37.2%) and the. On average five people – including children and adults - live in each household and the median per-capita family monthly income available is 96.5 US$.

**Table 3.**
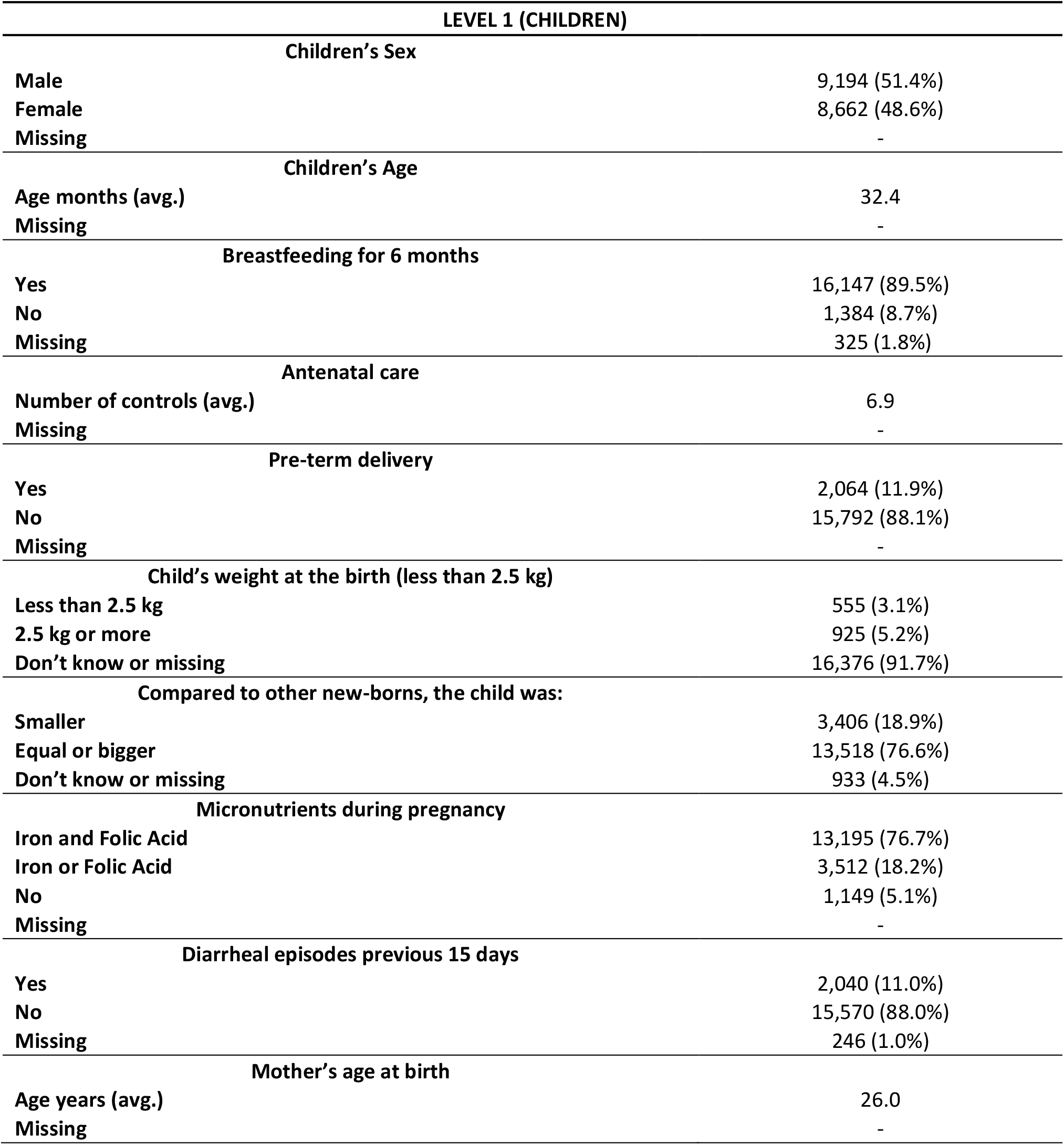
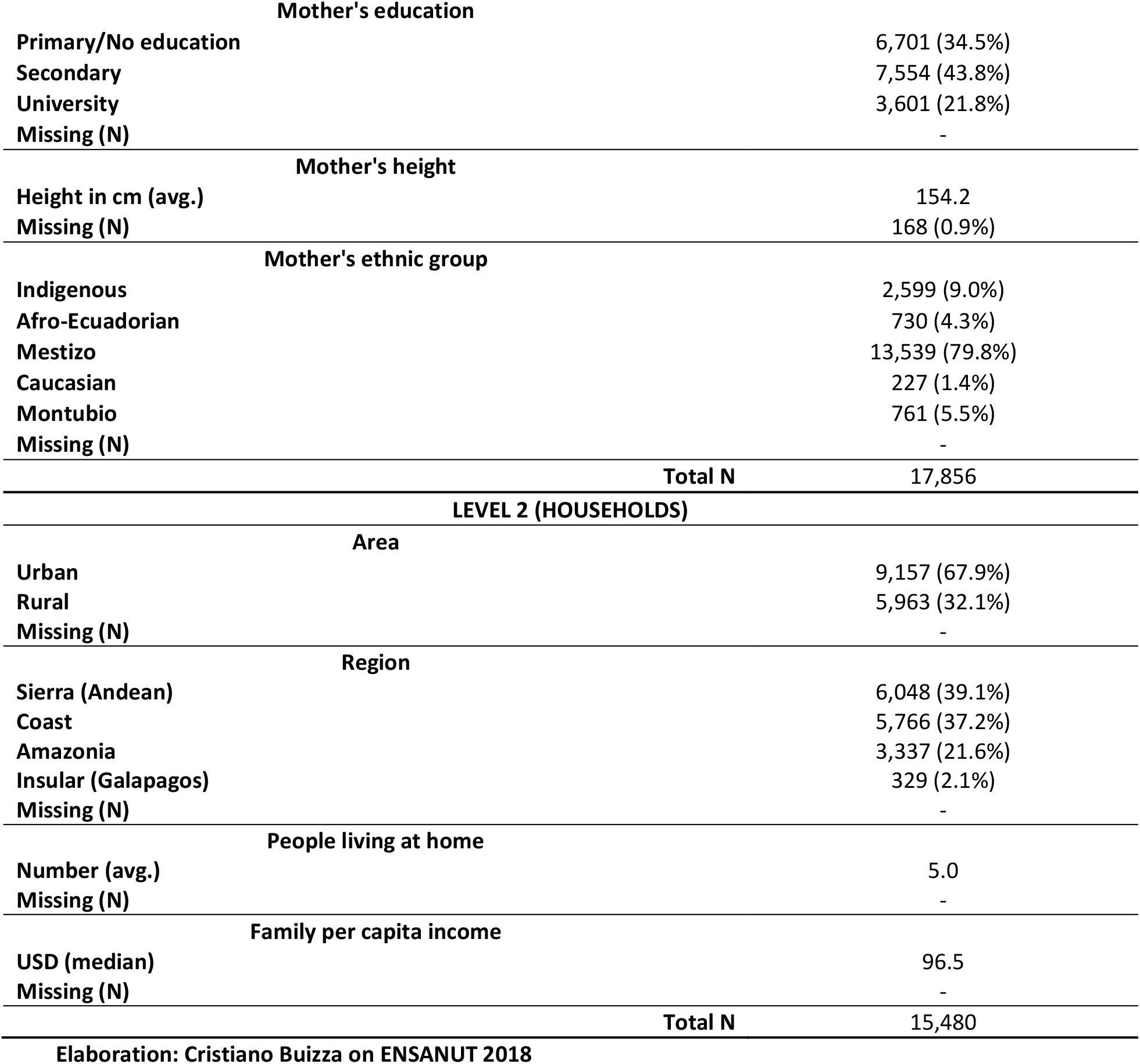
Socio-demographic and economic characteristics of the sample. N of the original sample and percentage after “svy”. Ecuador, ENSANUT 2018. Children with data on weight and height.

### 4.2 Variance Partition Coefficient and bivariate models

The second step of the analysis was the development of bivariate models to evaluate the strength and significance of the association of each exposure and each control variable with the outcomes. Before focusing on the crude effect of each factor on each response variable, the analysis of the proportion of the response variance that lies at each level of the null model is developed (Table4_i). The variance partition coefficient is estimated for each response variable. The formula adopted is 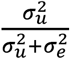, where 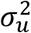 is the variance at household level and 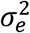 is the variance at individual level. In the case of logistic model, the variance at individual level is fixed and equal to 3.29, so in the case of stunting 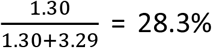 of the residual variation in the risk of stunting is attributable to unobserved household characteristics (Table4_iA). In the case of underweight 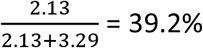 of the residual variation in the risk of being underweight is attributable to unobserved household characteristics (Table4_iB) and in the case of wasting 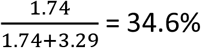 of the residual variation in the risk of wasting is attributable to unobserved household characteristics (Table4_i).

**Table 4.**
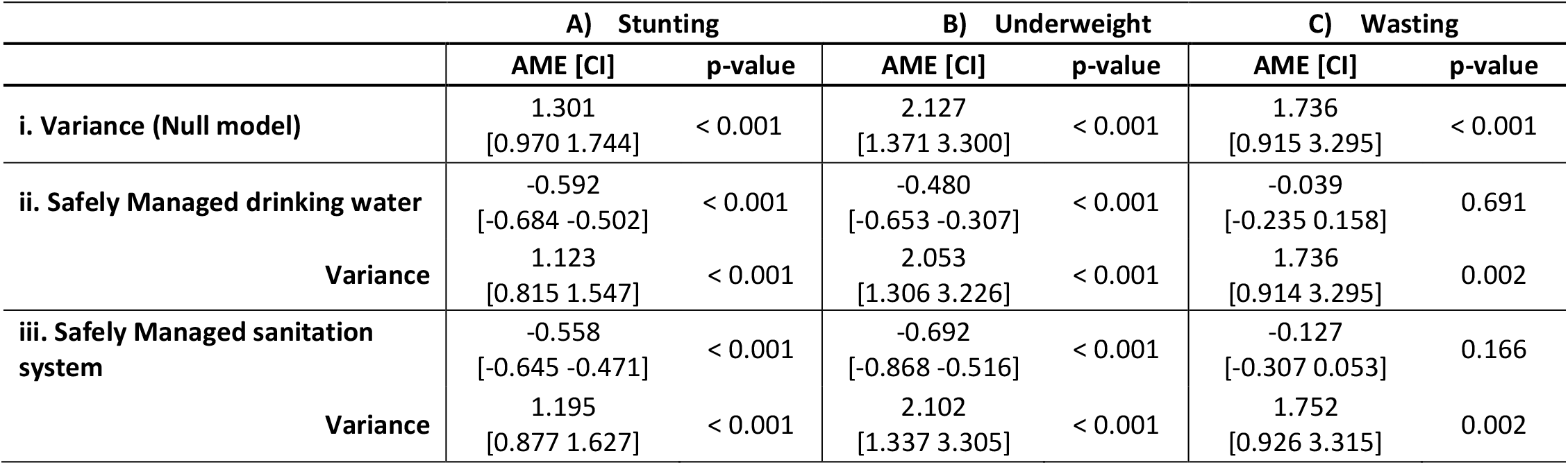
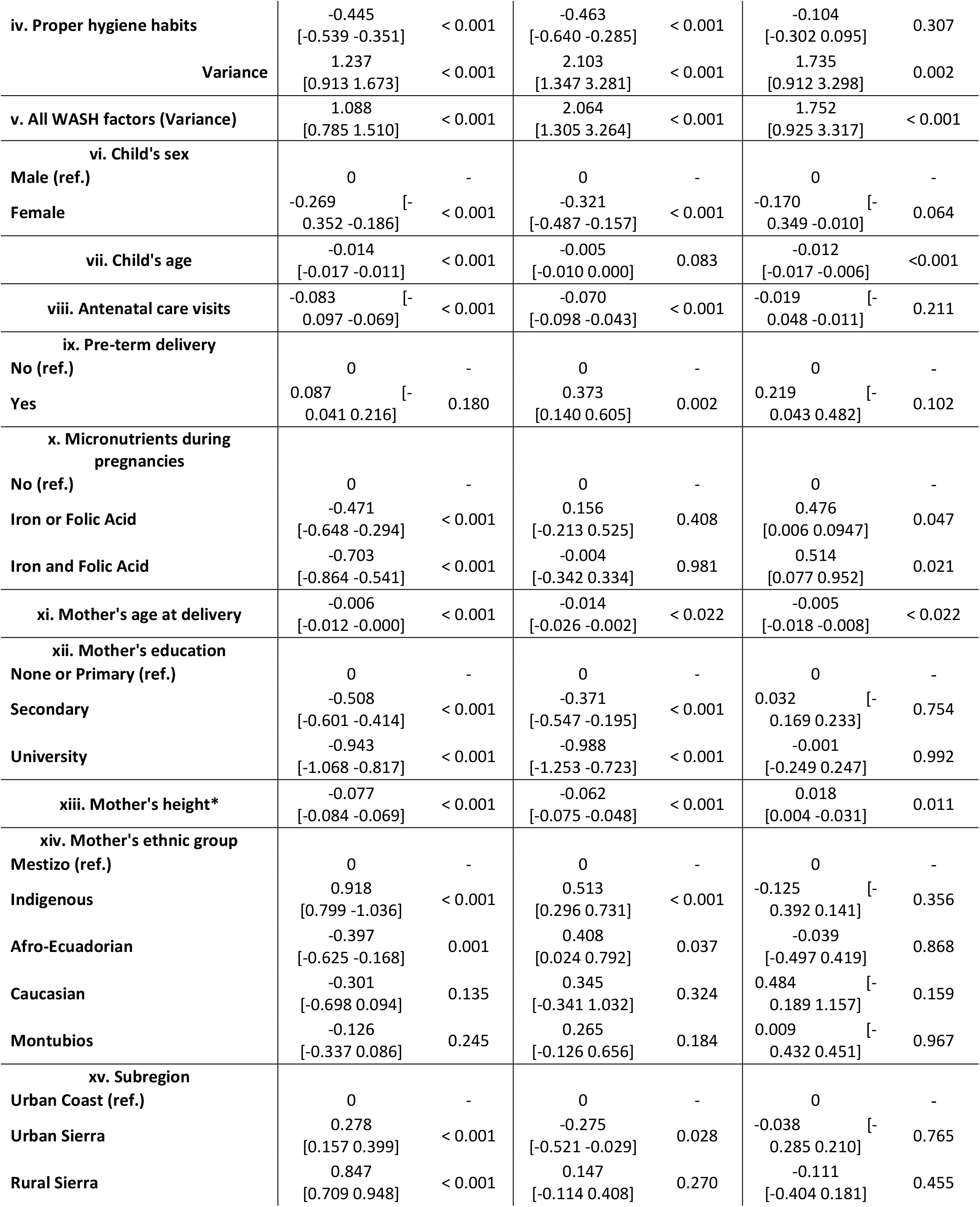
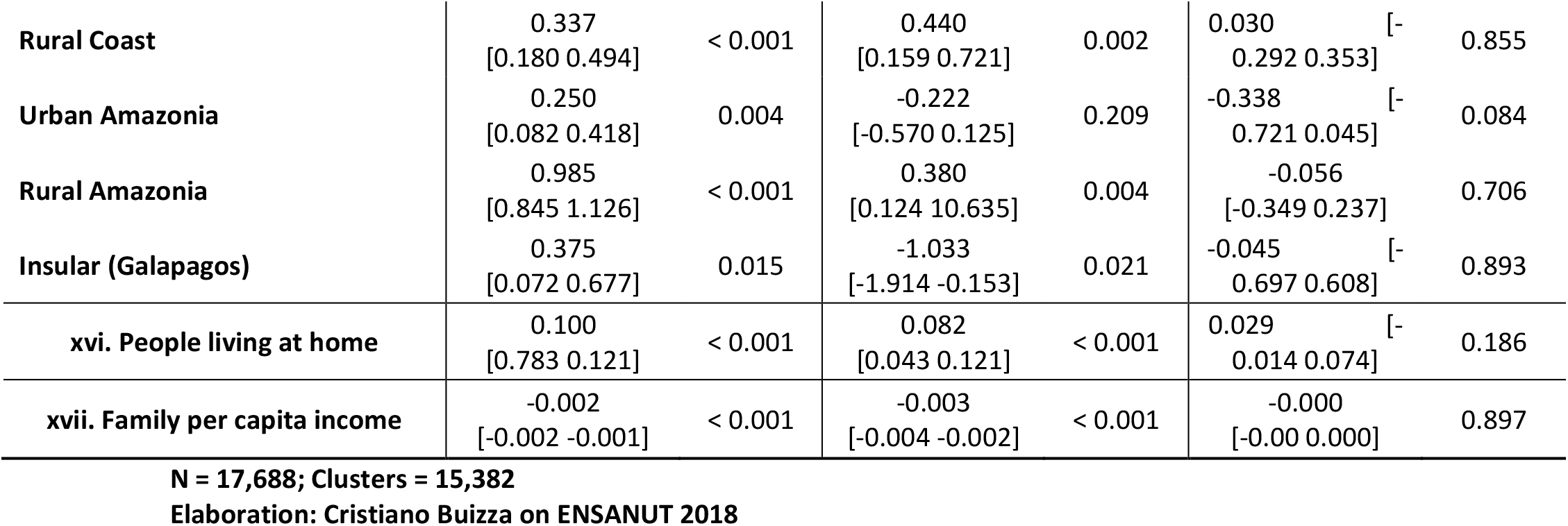
Average marginal effects (AME) of the two-level random intercept logistic bivariate association between the condition of stunting, underweight and wasting and each exposure and control variable. Children 6/59. Ecuador, ENSANUT 2018.

Table4 shows the crude effect of each variable on the three response variables separately. To avoid the effect of using a different number of observations, just children with complete information on all the variables were used (17,688 cases as the variable mother’s height is characterized by 168 missing data). Bivariate models between the exposures and the response variable also estimate the change on the variance. Focusing on the crude average marginal effects (AME) of stunting among children, a safe access to drinking water (Table4_iiA). denotes an average significant protective effect of -59.2% [95% CI -50.2% and -68.4%]. After controlling for this exposure, the residual response variation between cluster declines of 10.2% corroborating the importance of considering the factor to explain differences of stunting in Ecuador^12^. Similar effect is observed for safely managed sanitation (Table4_iiiA) with an average protective effect equal to -55.8% [95% CI -47.1% and -64.5%] and an average reduction of residual response variation of 6.4%. A lower but still important effect on stunting is observed adopting proper hygiene habits (Table4_ivA) with a protective effect equal to -44.5% [95% CI -35.1% and -53.9%]. Considering the joint effect of WASH factors, the residual stunting variation between cluster declines on average of 16.4% suggesting that pathogens play an important role on the short height of children in Ecuador (Table4_vA).

In the case of underweight, a safe access to drinking water (Table4_iiB) denotes an average significant protective effect of -48.0% [95% CI -30.7% and -65.3%]. The most important protective effect is observed if the household is equipped with a safe sanitation system (Table4_iiiB) when the probability of being underweight is reduced by 69.2% [95% CI -51.6% and -86.8%]. The implementation of proper hygiene habits (Table4_ivB) shows a lower but still important protective effect with an average reduction around -46.3% [95% CI -28.5% and -64.0%]. Considering the joint effect of WASH factors, the residual underweight variation between cluster declines of 2.9% suggesting that pathogens are not so important to define the weight of children in Ecuador (Table4_vB).

The comment on the effect of each confounder was developed after the final multivariable model when the effect of all the variables is considered jointly. The only aspect stressed in these initial bivariate analyses is the high importance both of hygienical factors (previously observed) and genetical/cultural^13^ factors on height. In fact, if minorities such as Indigenous and Afro-Ecuadorian (which normally show higher risk for poverty) suffer from underweight at a higher rate compared with Mestizos (on average +51.3% and +40.8%, Table4_xivB), the association is different focusing on stunting. Based on height (Table4_xivA), undernutrition is affecting just indigenous (+91.8%) and it is almost inexistent among Afro-Ecuadorian (−39.7%) compared with Mestizos. At the same time, every additional centimetre of the mother’s height from the average value (154.2 cm) reduces on average of 7.7% [95% CI -6.9% and -8,4%] the risk for the child of suffering from stunting (Table4_xiiiA). A similar effect is observed also for the underweight (−6.2%, Table4_xiiiB).

Different drivers seem to be associated with wasting. In this case, both the exposures and many of the confounders applied are unable to explain the variability of the phenomenon in Ecuador. The absence of common causes for stunting and wasting could corroborate the idea that undernutrition in Ecuador is not an issue depending on lack of food supplements but associated with other factors [Briend et al. 2015]. Due to the absence of significant bivariate association, wasting is not considered in the following part of the study.

### 4.3 Missing pattern

After having defined the potential variables to be considered, it was important to analyse the underlying mechanism which determines missingness. In fact, if the pattern of missing information is not completely at random or at least at random adding the confounder with the missing information to the model will affect the reliability of the outcomes estimated.

As the missingness on stunting, westing and underweight is associated with the same observations, only one multivariable model was developed to check for missing patterns. A binary variable equal to 1 was created if data are missing and 0 if not and the association with all the factors is observed applying a logistic model also considering the clustering effect at household level. As observed in Table5, no response variable and exposure are associated with the missing pattern of the mother’s height corroborating that missingness is at random. Same result is detected for confounders. The only significant exception is observed for the urban and rural Amazonia where data are more complete showing a lower risk for missingness. As the missingness seems to be non-random just within specific territories, sub-region variable will be added to the model in any case. Focusing on missingness on the response variables, outcomes do not show any substantial difference among exposures. Once again there is evidence against some areas of residence being missing at random as both urban and rural Coast shows a lower risk of missingness compared all the others sub-regions of the country. It is also observed a weak association between the risk of missingness and the age of the child, with younger children (6/23 months) characterized by a slightly higher risk of missing information on height and weight.

**Table 5.**
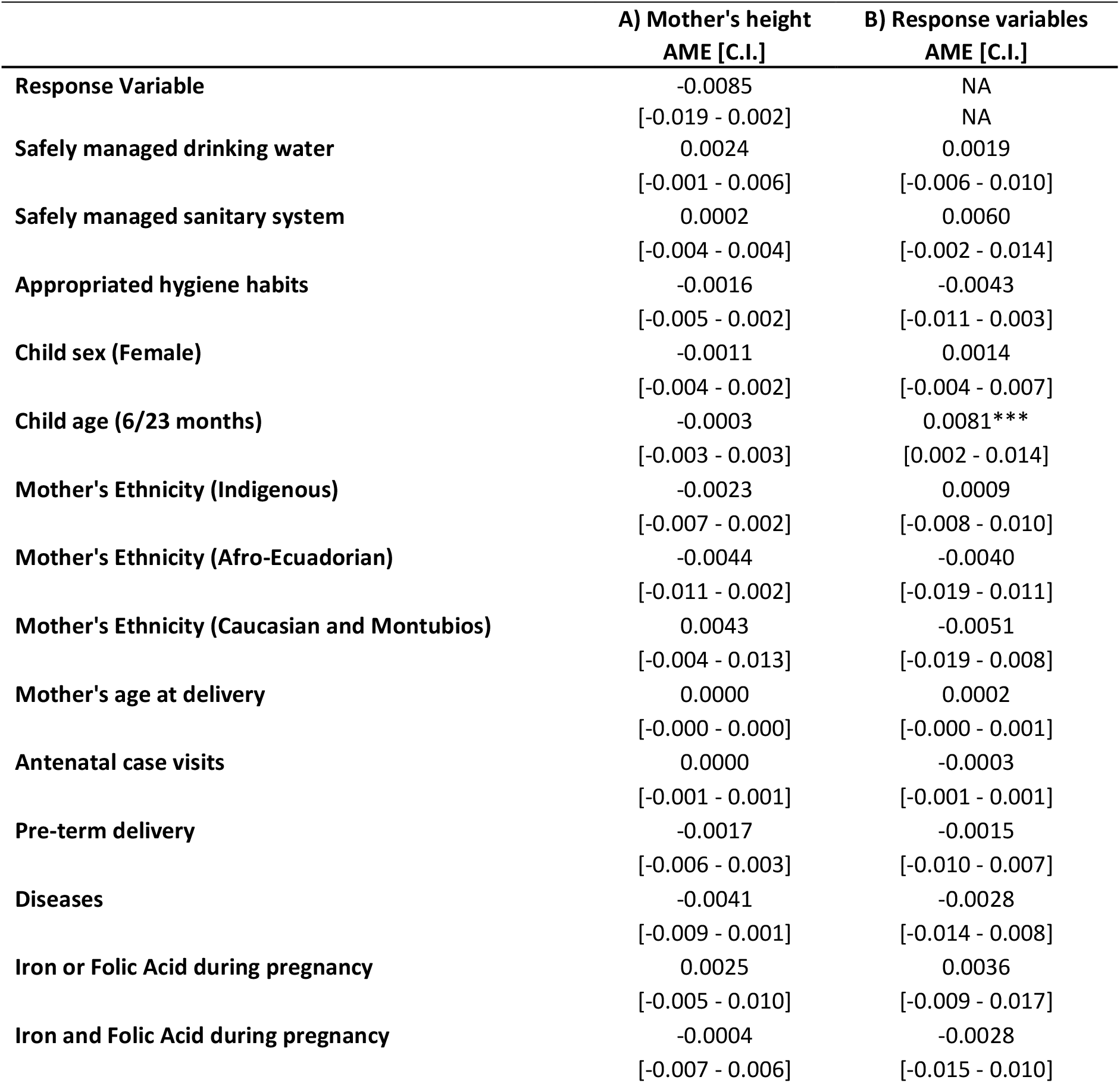
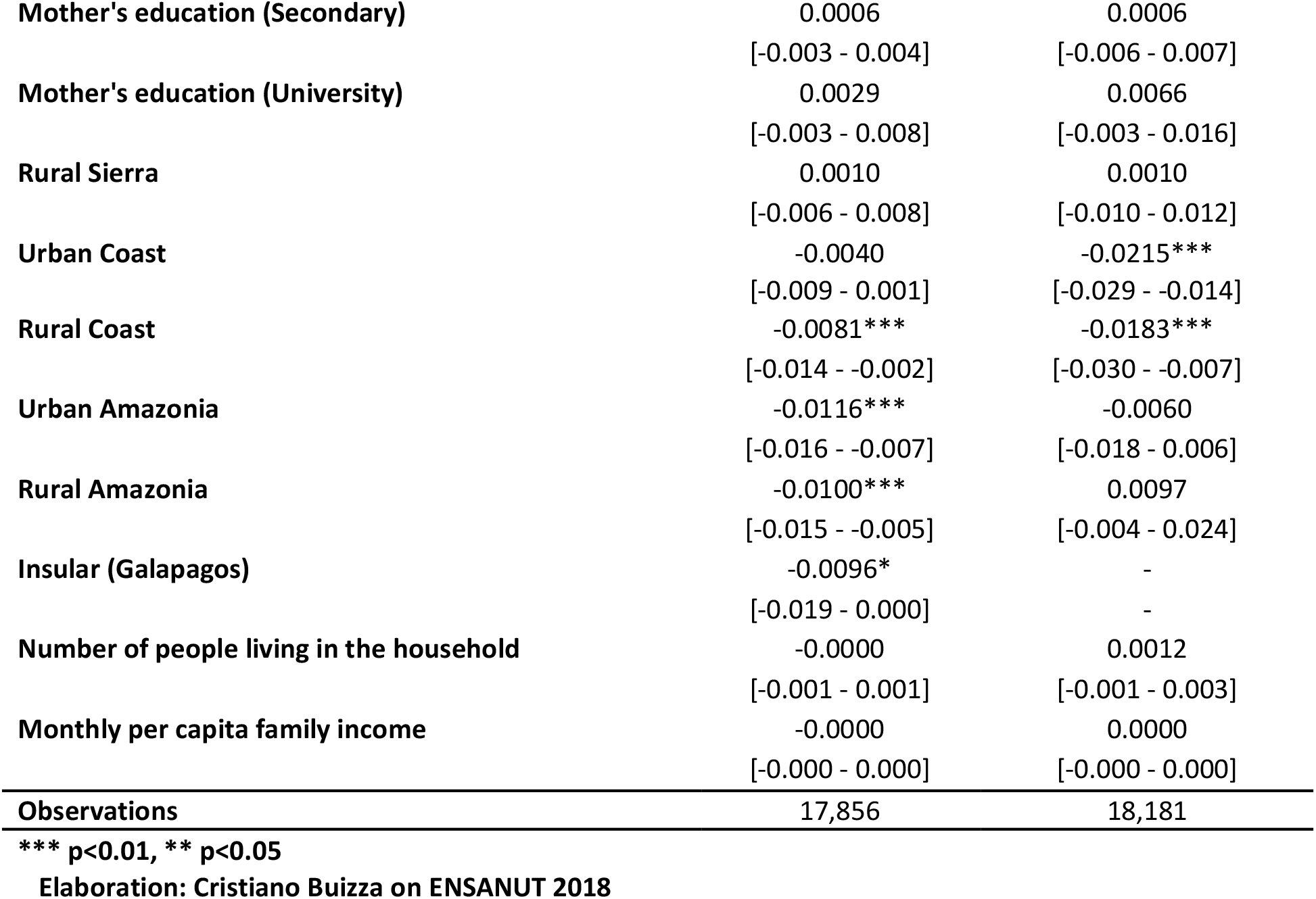
Average marginal effect (AME) of observing missing values on A) Mother’s height and B) Response variables. Parameters and confidence intervals estimated with a logistic model controlling for clustering at household level. Ecuador, ENSANUT 2018.

### 4.4 Multivariable logistic model with random intercept effects

The final models developed were multivariable logistic model with random intercept effects for stunting and underweight (Table6). In this case, the adjusted average marginal effects (AME) are provided. The 95% confidence interval (CI) of each parameter is included in the brackets. Adding confounders reduced the effect of the exposures for both stunting and underweight suggesting that their distribution is correlated to other factors. For both phenomenon a safely managed sanitation system shows the strongest protective effect with a reduction of 38.1% of the risk of being underweight [95% CI -16.9% and -59.4%] and a reduction of 14.9% of being stunted [95% CI -4.7% and -25.1%] (Table6_ii). Important protective effects against stunting are observed also thanks to safely managed drinking water [-10.9%, with a 95% CI suggesting a potential effect that can reach -21.6%] (Table6_i) and applying proper hygiene habits [-9.5%, with a 95% CI suggesting a potential protective effect of -19%] (Table6_iii).

**Table 6.**
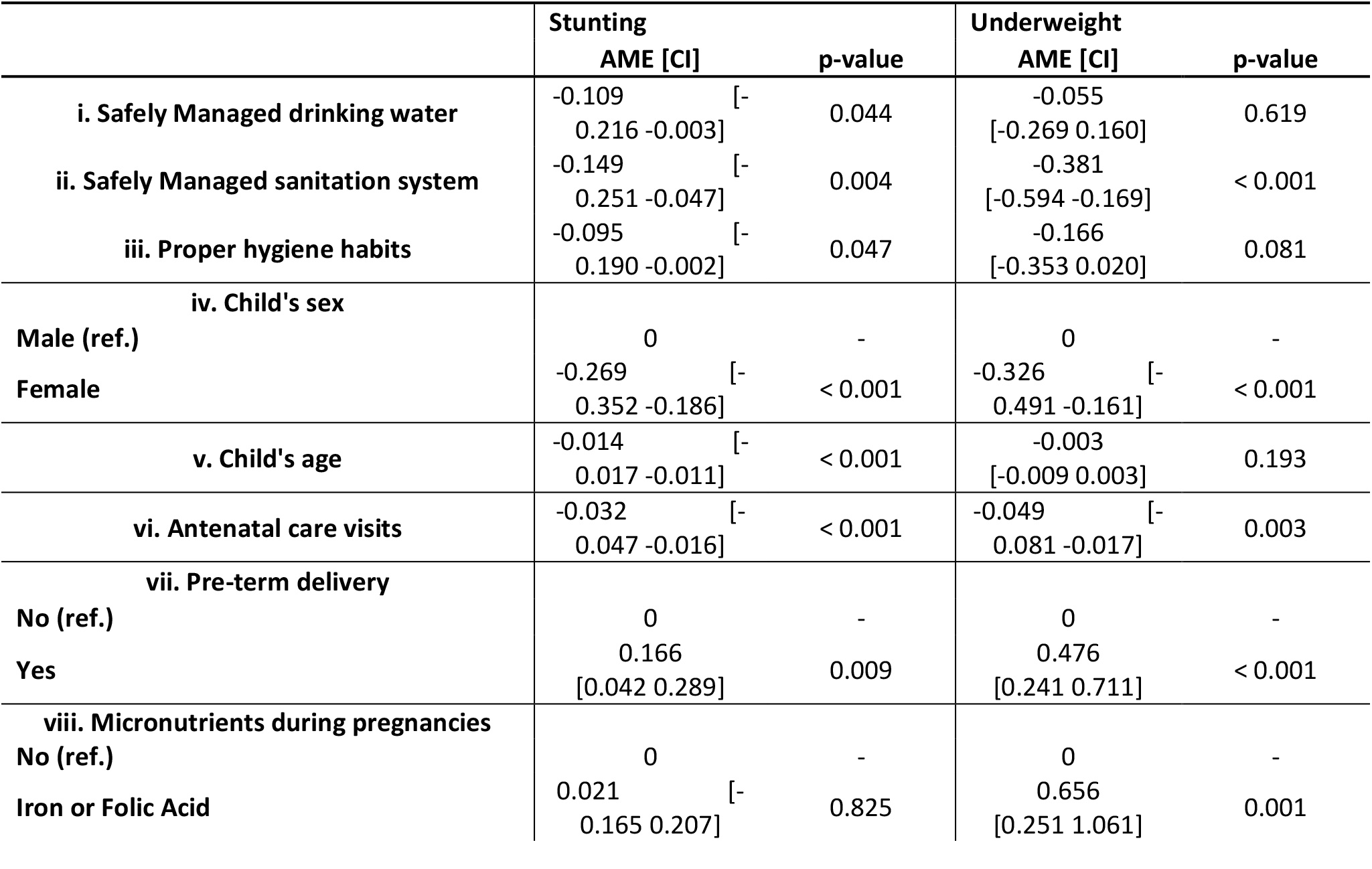
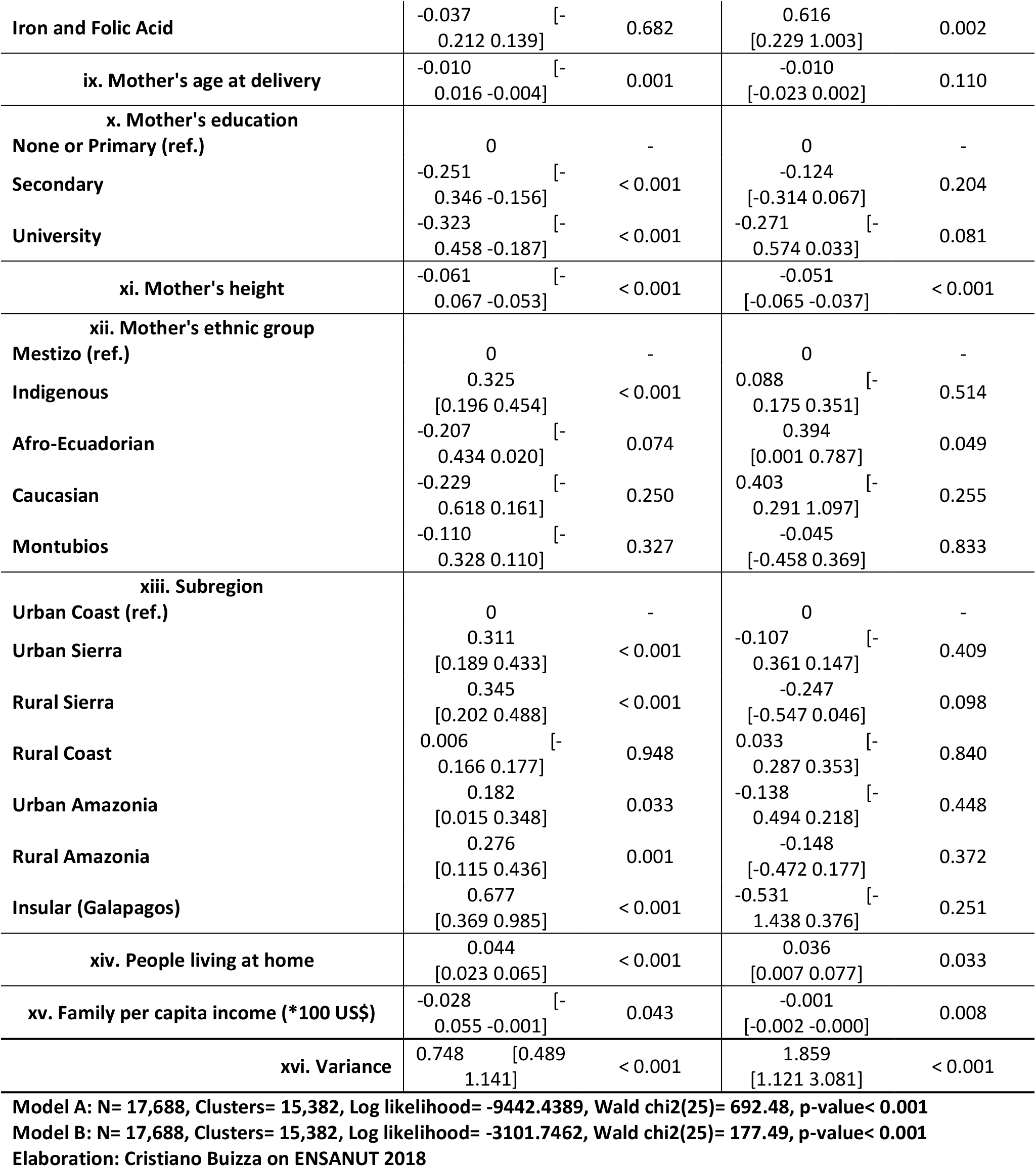
Average marginal effects (AME) of a two-level random intercept multivariable logistic model between the condition of stunting and underweight and the joint effect of exposures and control variables. Children 6/59 months. Ecuador, ENSANUT 2018.

As shown in Table6_iv, in both cases, female population is characterized by an important and significant lower risk for both stunting (on average -26.9%) and underweight (on average -32.6%) suggesting that boys are more exposed to health inequalities than women in the same age groups as observed in Wamani et al. (2007). Age pattern (Table6_v) confirms that stunting is lower after the 24^th^ month of life while no change is observed for underweight. A pre-term delivery (Table6_vii) shows a strong growth of the risk for both stunting (16.6%, 95% CI 4.2%-28.9%) and especially for underweight (47.6%, 95% CI 24.1%-71.1%).

Several antenatal care visits higher than the minimum required (equal to 4) reduces on average the risk of stunting of 3.2% and the risk of underweight of 4.9% for each additional visit suggesting that malnutrition is a phenomenon that partially can be avoided if it receives an early treatment (Table6_vi). A higher mother’s education (Table6_x), which is strictly related with wealth and healthier lifestyle, significantly reduces the risk for stunting (−32.3%, 95% CI -18.7% and -45.8%) and partially for underweight (on average -27.1%, although the 95% CI is slightly inconclusive).

As previously stressed, in Ecuador stunting seems to be associated with genetical and cultural aspects represented by mother’s ethnic group (Table6_xii) and mother’s height (Table6_xi). It is important to stress the substantial change of the adjusted AME of ethnic group compared with the crude AME previously estimated (Table4) which suggests that the distribution of this factor is strongly associated with other variables. The variable representing genetics factors such as mother’s height (Table6_xi) confirm the strong preventive action against stunting and underweight observed in Table4.

For each additional person living in the household (Table6_xiv), used as a proxy for the number of people sharing WASH facilities, the risk for suffering from stunting is 4.4% higher [95% CI 2.3%-6.5%] while the risk of underweight is 3.6% higher [95% CI -0.7%-7.7%]. Also, the level of household wellbeing shows an important effect (Table6_xv). Per each 100 US$ additional to the median family per capita income the risk for stunting decreases on average of -2.8%.

Focusing on the residual response variance still existing after applying all the variables, the model for stunting denotes a drop of 34.6% compared to the null model denoting a good ability to explain response variability (Table6_xvi). The assumption of Normality of level-2 residuals seems to be respected with a Skewness of 0.979 and kurtosis of 3.035. In the case of the model for underweight the multivariable model obtained a reduction of the residual response variance of just 8.5%. In this case, some problems are observed for the assumption of Normal distribution with a Skewness of 4.020 and Kurtosis of 18.823. In both cases, the test at the end of the table6 corroborates the importance of considering a correlated model instead of a model assuming independency of observations.

## 5. DISCUSSION

### 5.1 Summary

Results indicate that WASH factors play a fundamental role in preventing the risk for chronic undernutrition (stunting) and partially in preventing underweight. The strongest protective effect is observed when sanitation system is managed safely which reduces the risk of stunting of 14.9% and the risk of being underweight of 38.1% (Table6_ii). This outcome leads to the idea that a massive investment is necessary as just 53.9% of households (Table2) can be classified as completely safe managed in sanitation system. This is an important finding as it also suggests that the way traditionally adopted to classify household as safe or unsafe probably is not appropriated to capture completely the phenomenon. The preventive effect associated with underweight (−38.1%) is among the highest observed in literature. This is probably associated with the fact that in this analysis it was impossible to control for dietary aspects which could have inflated the effect of the exposure. Also the application of proper hygiene habits (use of water and soap) denotes some protective effects for both phenomenon: on average - 9.5% for stunting and -16.6% for underweight (Table6_iii). A safely managed drinking water shows a clear impact just on chronic undernutrition (−10.9%) while no clear association is observed for the risk of being underweight (Table6_i). No association has been observed with acute undernutrition (wasting). In general, as observed in literature [Dangour et al. 2013, Luby et al. 2018, Null et al. 2018, Bekele et al. 2020] also in Ecuador the sanitary system seems to play the most important role in preventing child growth failure.

Genetical and cultural factors such as mother’s height and ethnic group show a strict relationship with stunting. At the same time, the initial indicators show that in Ecuador the prevalence of wasting and undernutrition is uncommon. This is an important aspect to consider as it can suggest that developing public policies focused on the provision of food program it could not to be useful to curb chronic undernutrition in Ecuador. Once again, to corroborate this result, models should incorporate variables related to dietary habits to evaluate if effects observed for ethnic groups depend on different dietary habits.

Although genetical and cultural aspects can play an important role, this does not mean that undernutrition is something predestined and external to human action. Adopting proper lifestyles and attending early and regularly a doctor during pregnancy reduce the risk of premature delivery, which is associated with a robust higher risk of stunting (+16.6%) and underweight (+47.6%) (Table6_vii). A proactive strategy based on early diagnosis and a consequent early treatment are fundamental steps to reduce the risks. A number of antenatal care visits higher than the minimum required reduces the risk of stunting of 3.2% and the risk of underweight of 4.9% for each additional visit (Table6_vi). Educated women are another aspect able to curb children undernutrition. The improvement of the level of education of the mothers significantly reduces the risk for stunting (−32.3%) and partially the risk of being underweight (−27.1%) (Table6_x). Overcrowded household and poverty increase the risk of stunting and underweight.

### 5.2 Limitations and recommendations

The strategy adopted in this study is based on the research of common drivers to explain the variability of stunting, underweight and wasting, but a certain improvement could require the definition of specific factors for each item. This issue seems to affect principally the phenomenon of wasting which is a combination of both height and weight.

Several issues that could threaten the internal validity are observed. The survey is generally based on self-reported data which could produce differential misclassifications if mothers affected by stunting, underweight and/or wasting are characterized by a higher level of accuracy or interest in recalling and providing data. Moreover, the source of information is mixed as the height and the weight and the aspects associated with proper hygiene habits are measured or corroborated by the interviewer, while all the other information derived by subjective answers. This issue normally increases the risk for biased estimations. To minimize the risk for recall bias a subsample of the last three children under-5 born in the five-years period before the interview has been used.

Cross-sectional data are characterized by risks for reverse-causality. This issue seems to affect the association between micronutrients assumption during pregnancy and the outcomes with a higher risk of underweight among mothers who have received iron and/or folic acid. As previously stressed, this could be the consequence of the fact that mothers who discovered that their children were underweight during pregnancy started to use micronutrients. In general, to avoid this problem, confounders associated with events occurred after the delivery such as the number and type of post-delivery visits were not included in the model as they could be the consequence of the observed outcomes. Additionally, cross-sectional data uses the prevalence – and not the incidence - of phenomenon generating limitation to test hypothesis

No information on dietary habits is included in the model as no exhaustive data are provided for the last three children which are used in this study. Therefore, future analysis in Ecuador should be focused just on the last child born as more information are collected on this fundamental topic. However, adding more variables should be done taking care of the potential loss of information generated by missing data which are affecting alternative variables not included in the current study.

## Data Availability

All data produced are available online at
https://www.ecuadorencifras.gob.ec/salud-salud-reproductiva-y-nutricion/

## Footnotes

2 A detailed explanation is provided in the methodological paragraph.

3 Implausible values are eliminated when the value calculated is below or above -6SD/+5SD compared to the median of the reference population in the case of weight-for-age, and - 6SD/+6SD compared to the median of the reference population in the case of height-for-age.

4 Adding people which cleaned just sometimes septic tanks or pit latrine (another improper way to avoid pathogens) the percentage is almost 100%.

5 Due to the extremely small amount of people declaring using surface water, this type of water is included in unimproved category and not considered as a third category.

6 In the case of the variables related to breastfeeding habits and diarrheal episodes, although the amount of missing data is not high, the logistical model developed shows strong evidence against a missing at random mechanism for several variables.

7 Outcomes of figure1 and Table1, Table2, Table3 and Table4 are obtained considering the complex structure of the survey.

8 The overall prevalence of stunting among all the under-five children is 23.8% which is slightly higher than the official estimate provided by the Ecuadorian National Institute of Statistics which is 23.1%.

9 The overall prevalence of underweight among all the under-five children is 4.9% which is slightly lower than the official estimate provided by the Ecuadorian National Institute of Statistics which is 5.2%.

10 The overall prevalence of wasting among all the under-five children is 3.7% as the official estimate provided by the Ecuadorian National Institute of Statistics.

11 From now on, “children” would be used to represent “children 6/59 months”.

12 If in the null model 28.3% of the residual variation in the risk of stunting is attributable to unobserved household characteristics now is 1.123/ (1.123+3.29) = 25.4% which is 10.2% less. The same strategy is applied to estimate all the other reductions of residual variance.

13 Effect of ethnic group could also represent the effect of a different diet or cultural aspects which are not considered in this study.

